# AGO2 protein: A Key Enzyme in the miRNA Pathway as a Diagnostic and Prognostic Biomarker in Adrenocortical Carcinoma

**DOI:** 10.1101/2024.02.19.24302333

**Authors:** Anila Hashmi, Gyorgy Hutvagner, Stan Sidhu, Alexander Papachristos

## Abstract

**Context:** Adrenocortical carcinoma (ACC) is a rare and aggressive malignancy. Current treatment algorithms are associated with diagnostic limitations, high recurrence rates and poor prognosis. Identifying specific biomarkers that facilitate accurate diagnosis and provide prognostic insights could significantly enhance the patient outcomes in ACC.

**Objective:** To investigate whether microRNA machinery, specifically argonaute 2 (AGO2), a key enzyme in the miRNA pathway, has the potential to be a diagnostic and prognostic biomarker for adrenocortical carcinoma (ACC).

**Design:** This study analyzed mRNA expression of genes involved in the miRNA biogenesis pathway using RNASeq data from The Cancer Genome Atlas (TCGA) and The Genotype-Tissue Expression (GTEx) dataset, followed by target protein quantification in tissue samples using commercial ELISA kits.

**Setting:** Publicly available mRNASeq datasets (TCGA-GTEX) and frozen tissue samples from the tumour bank of the Kolling Institute of Medical Research.

**Participants:** We analyzed data for 79 ACC and 190 normal adrenal cortex (NAC) samples from the TCGA and GTEx datasets, as well as for 31 other cancer types from the TCGA. We then performed protein quantification in 15 NAC, 15 benign adrenal adenoma (AA), and 15 ACC tissue homogenates.

**Intervention(s):** None.

**Main Outcome Measures:** AGO2 mRNA and protein expression in ACC and its prognostic correlation.

**Results:** AGO2 was significantly overexpressed in ACC, compared to NAC and AA (p<0.001). Kaplan– Meier survival analysis revealed that higher expression of AGO2 was associated with significantly worse overall survival in ACC (HR 7.07, p<0.001). Among all 32 cancer types in TCGA, AGO2’s prognostic utility was most significant in ACC.

**Conclusions:** AGO2 holds potential as a diagnostic and distinct prognostic biomarker in ACC.

## Introduction

Adrenocortical carcinoma (ACC) is a rare and highly aggressive malignancy of the adrenal gland. Five-year survival rates vary based on disease stage at diagnosis, ranging from 60-80% for localized tumours to 0-28% for metastatic disease (1). Currently, surgical resection remains the only curative therapeutic option. For unresectable disease, systemic therapy is recommended by clinical practice guidelines, however, the efficacy of these treatments is limited, with objective response rates of less than 25%, and significant side-effects(1,2). Even after curative resection, disease recurrence occurs in more than 60% of patients, and poses a significant therapeutic challenge (3). Moreover, despite advances in the genomic characterisation of ACC (4,5), there are currently no biomarkers that facilitate diagnosis, pathological prognostication, or monitoring for recurrent disease after curative resection(6–8).

MicroRNAs (miRNAs) are small non-coding RNAs that regulate more than 60% of protein coding genes by interacting with messenger RNA (mRNA) (9). The differential expression of miRNAs between ACC and adrenal adenoma has recently emerged as a potential diagnostic and prognostic indicator. Specific miRNAs, such as the upregulation of miR-503, miR-210, miR-483-5p, and miR-483-3p, and the downregulation of miR-195, miR-497, and miR-335, have been identified as potential markers for ACC (10). However, the lack of significantly different expression of hsa-miR-483-3p and hsa-miR-483-5p between adrenal myelolipoma and ACC limits their clinical utility (11). Furthermore, conflicting patterns of miRNA expression in ACC and adrenocortical adenoma (AA), have been reported (12,13). These discrepancies highlight the complexity of miRNA regulation in ACC and the need for standardized quantification protocols and rigorous validation. Currently, the utility of miRNAs as biomarkers is limited by their low expressed concentrations, lack of standardised analytical methodologies and lack of specificity to tumour types(14).

Advances in RNA sequencing technology have facilitated the identification of miRNA isoforms (isomiRs) that may have clinical utility in the context of ACC. The miRNA biogenesis pathway involves a series of tightly regulated interdependent steps, starting with the transcription of primary miRNAs (pri-miRNAs)(15),which are cleaved into precursor miRNAs (pre-miRNAs) by the Drosha-DGCR8 complex(16). The pre-miRNAs exported to the cytoplasm by Exportin-5 and RANGTP(17,18), where they are further processed by the TARBP2 and Dicer enzyme into miRNAs(19–21). The miRNA duplexes are incorporated into one of the Argonaute proteins that unwinds the double stranded miRNA. One strand of the miRNA becomes the part of the RNA-induced silencing complex (RISC) while the other strand is degraded. The RISC complex inhibits mRNA translation and gene expression (22–25) (Fig1) Therefore, changes in miRNA expression may alter gene expression in a manner that leads to tumour development (26).

In various cancers, such as clear cell renal carcinoma (27), ovarian carcinoma (28), leiomyosarcomas (29), and breast cancer (30), the deregulation of miRNA-processing complexes has been observed, indicating their potential role in tumorigenesis. In ACC, two notable studies have reported contrasting findings. Caramuta et al. discovered significant gene and protein overexpression of TARBP2, DICER1, and DROSHA in ACC compared to AA and NAC, with TARBP2 overexpression found to be the dominant differentiator between ACAs and ACCs (31). In contrast, de Sousa et al. reported no significant differences in TARBP2 gene or protein levels between ACCs and ACAs, instead finding weak Dicer1 expression was associated with reduced survival in metastatic ACC (32). In this study, we focus on understanding the role of the miRNA machinery components in adrenocortical carcinoma (ACC). AGO2 is a key component of the RNA-induced silencing complex (RISC) and guides miRNAs to their target genes, thereby regulating gene expression at the post-transcriptional level (33). Through a comprehensive analysis of AGO2 and related miRNA genes, we aim to explore their potential as novel pathological prognostic biomarkers for ACC.

## 2. Materials and Methods

### 2.1. RNASeq Data Analysis for miRNA Biogenesis Genes in ACC

We sourced RNASeq data from two public repositories: The Cancer Genome Atlas (TCGA) for cancer samples, and The Genotype-Tissue Expression (GTEx) project for normal tissue samples. TCGA, a collaborative program between the National Cancer Institute (NCI) and the National Human Genome Research Institute, has molecularly characterized primary tumours and matched normal tissue across 33 cancer types, providing a comprehensive platform for researchers to access and analyze cancer data. The GTEx project collects normal tissue samples and characterises tissue-specific gene expression and regulation using for molecular assays such as whole genome sequencing (WGS), whole exome sequencing (WES) and RNA-Seq.

Our bioinformatic analysis focused on the expression of core components in the miRNA biogenesis pathway, specifically AGO2, DGCR8, XPO5, RAN, DROSHA, DICER, and TARBP2, in adrenocortical carcinoma (ACC). Normalized RNA sequencing (RNA-seq) data specific to miRNA biogenesis genes for normal adrenal cortical tissue was obtained from the Genotype-Tissue Expression (GTEx) project, and from The Cancer Genome Atlas (TCGA) for adrenocortical carcinoma (ACC). The TNMplot bioinformatics web tool was used for data retrieval(34).

### 2.2. Survival analysis

Survival analysis paired gene expression data and survival data from The Cancer Genome Atlas (TCGA), using the Encyclopedia of RNA Interactomes (ENCORI) database (35). Kaplan-Meier survival analysis was performed on the UCSC Xena platform(36). To explore specificity of the prognostic value of AGO2 expression to ACC, and the potential interaction with other miRNA biogenesis genes, survival data for 32 different cancers was accessed from TCGA, including clinicopathological data where available.

### 2.3. Tumour Samples

The study received ethics approval from the Northern Sydney Local Health District Human Research Ethics Committee (2020/ETH01931). Tissue samples, including adrenocortical carcinoma (ACC), benign adrenocortical adenoma (AA), and normal adrenal cortex (NAC), were sourced from the Tumour Bank of the Kolling Institute of Medical Research. The Kolling Institute Tumour Bank Access Committee granted access to these samples (reference NETBMC #20-49). All participating patients provided informed consent for the use of their tissue samples and the collection of associated clinical data. At the time of adrenalectomy, tissue samples were immediately snap-frozen in liquid nitrogen and subsequently stored at −80°C. All ACC samples utilized in this study were histologically confirmed according to accepted diagnostic criteria (37).

### 2.4. Protein expression analysis

Snap-frozen tissue samples, including 15 NAC, 15 AA, and 15 ACC, were obtained from the Kolling Institute Tumour Bank. Tissue homogenates were prepared by washing the tissue with pre-cooled phosphate-buffered saline (PBS) buffer (0.01M, pH=7.4). The tissue samples were then homogenized in Lysing Matrix A tubes (MP Biomedicals, Australia). Homogenization was performed using the FastPrep-24™5G (MP Biomedicals) bead beating grinder and lysis system, according to the manufacturer’s guidelines. Protein expression levels of miRNA biogenesis genes were measured using Human Protein ELISA Kits according to the manufacturer’s instructions, and included AGO2, DGCR8, DROSHA, RAN, XPO5 (Abebio-Co. Ltd.) and TARBP2 and DICER1 (Fine Biotech Co., Ltd.). Protein concentrations were measured by comparing the optical density to standard controls using a microplate reader (TECAN Spark absorbance reader). (Figure 2).

**Figure 1.**
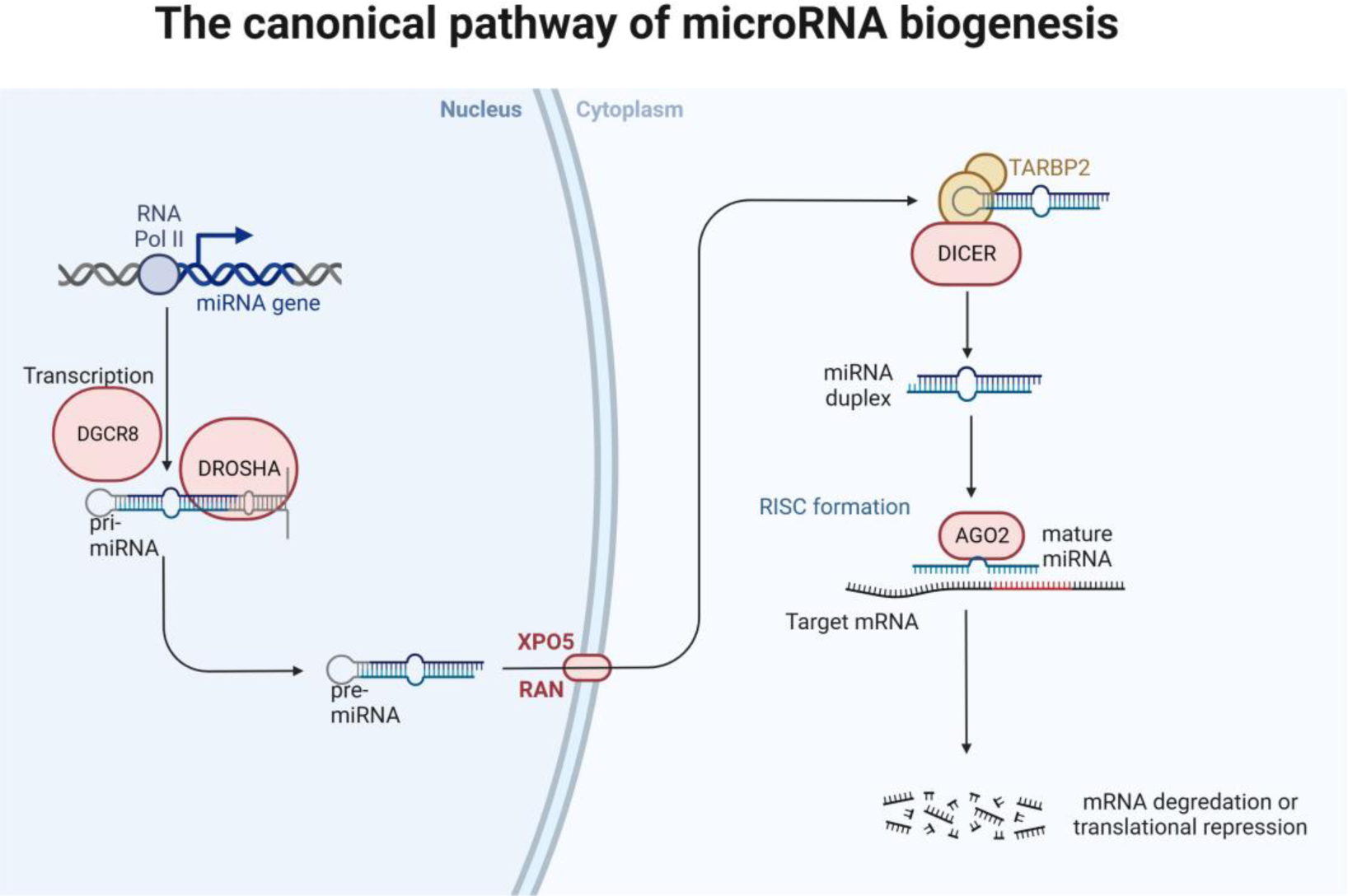
The Canonical pathway of microRNA biogenesis. MicroRNA (miRNA) genes are transcribed by RNA polymerase II (Pol II) to generate the primary microRNA (pri-miRNAs). Drosha/DGCR8 cleavage complex removes the tails of the pri-miRNA and form the shorter stem loop structure called precursor microRNA (pre-miRNA). Pre-miRNA exported from the nucleus to cytoplasm by Exportin-5 (XPO5) and its cofactor RAN. In the cytoplasm pre-microRNA released from exportin-5 and further processed by Dicer complex and TARBP2 to produce RNA duplex. The guide strand of the mature miRNA then incorporated into the RNA-induced silencing complex (RISC). Following the unwinding, microRNA guides RISC to conserved recognition sites in the target messenger RNA and inhibit its expression. Created with BioRender.com.

**Figure 2.**
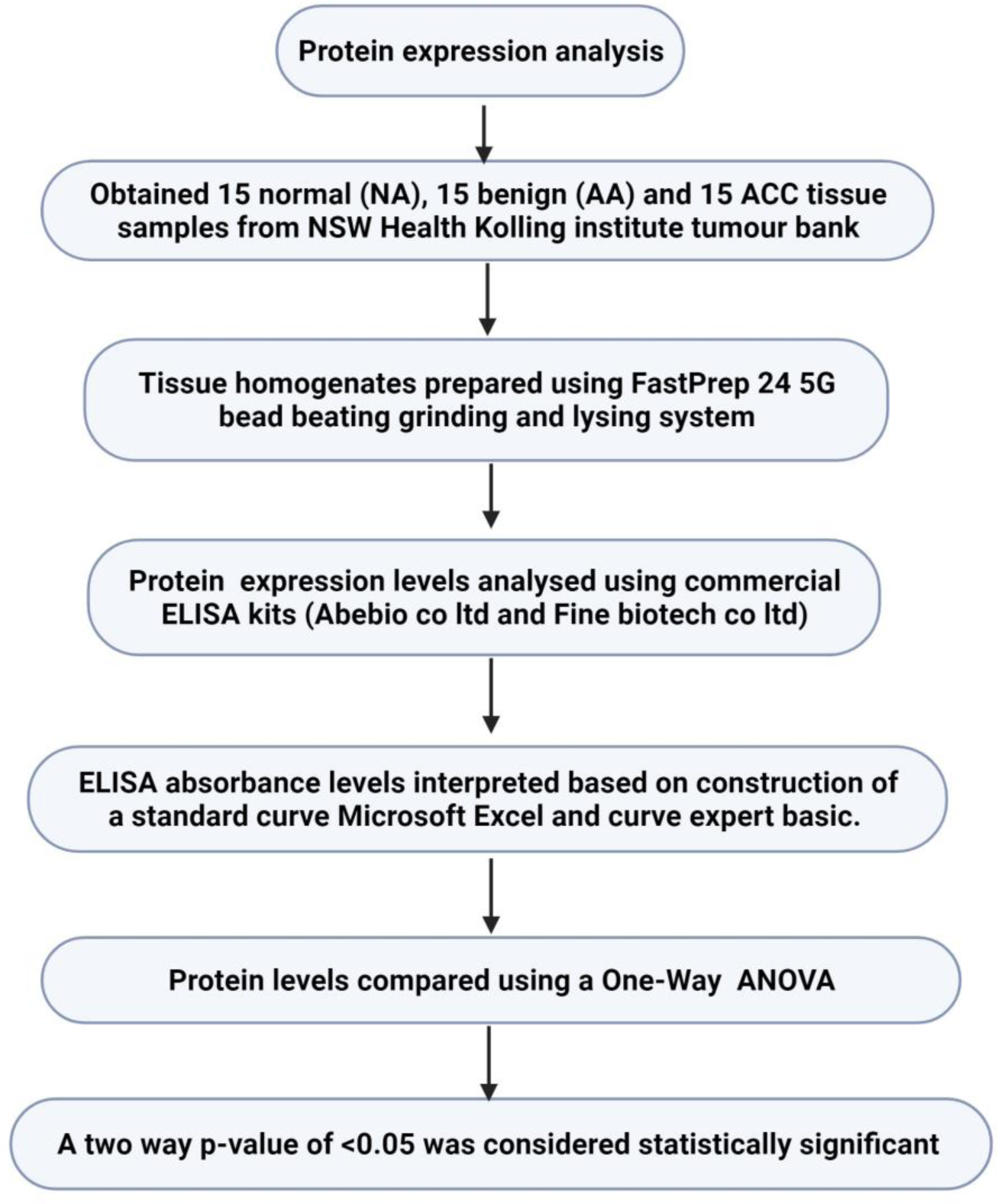
Flow chart of the protein expression analysis method. Created with BioRender.com

### 2.5. Statistical Analysis

Statistical analysis was performed using GraphPad Prism, Version 9 (GraphPad Software, CA, USA). For gene expression data analysis, a two-way Analysis of Variance (ANOVA) was employed to compare the expression levels between groups. The log-rank test was used to compare survival outcomes between groups; for both gene expression and gene survival analysis, a p-value of <0.05 was considered statistically significant. To explore the correlation between gene expression and tumour staging in ACC, a one-way ANOVA was utilized with a p-value threshold of < 0.05. ELISA absorbance levels were interpreted based on the construction of a standard curve in Microsoft Excel (Version 2306 Build 16.0.16529.20166) and Curve Expert Basic (V.1.4-USA), with protein levels compared using a one-way ANOVA and a p-value threshold of < 0.05. Additionally, the Receiver Operating Characteristic (ROC) curve was employed to determine the optimal cut-point for AGO2 protein levels, balancing sensitivity and specificity in the diagnosis of ACC.

## 3. Results

### 3.1. Analysis of miRNA biogenesis gene expression in adrenocortical carcinoma and normal adrenal cortex

In the RNA-seq data from the GTEx project and TCGA, AGO2, RAN, and TARBP2 were significantly upregulated in ACC samples compared to the normal adrenal cortex (p <= 0.001). Conversely, DGCR8 expression was slightly higher in the normal adrenal cortex than in ACC (p=0.014). No statistically significant differences were observed in the expression levels of DROSHA (p=0.24), DICER1 (p=0.19), and XPO5 (p=0.66) (Figure 3).

**Figure 3:**
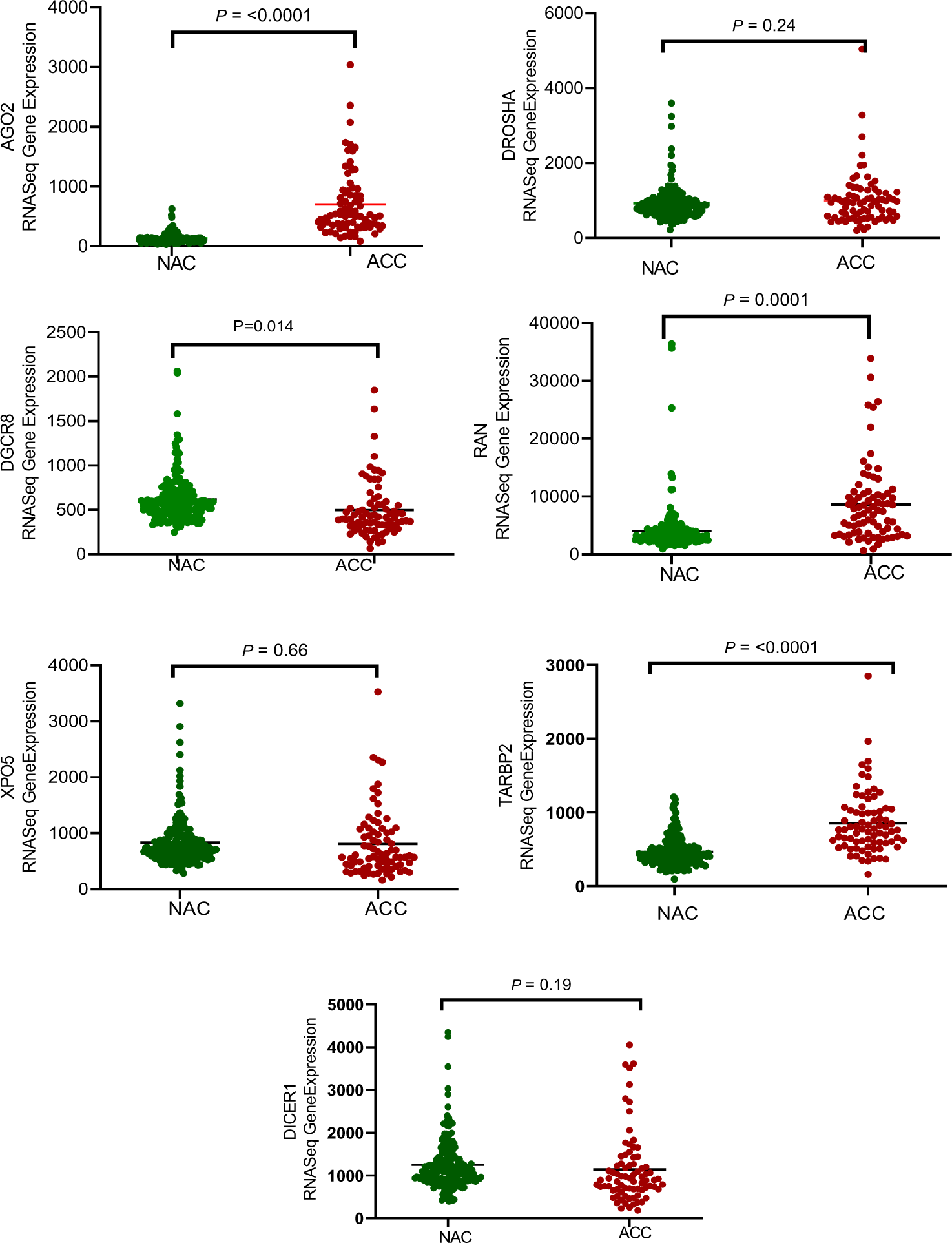
Gene Expression Analysis of miRNA Biogenesis Genes in Adrenocortical Carcinoma (ACC) and Normal Adrenal Cortex (NAC) tissue samples. The expression levels of miRNA biogenesis genes (AGO2, DROSHA, DICER1, DGCR8, XPO5, and RAN) were compared in 79 malignant ACC and 190 normal adrenal cortex tissue samples using RNA-seq data from the TCGA and GTEX datasets (34). Among these genes, AGO2 showed significantly higher expression in ACC samples than in normal samples (p<0.001), whereas normal samples displayed minimal or no expression of AGO2. Moreover, the upregulated gene expression of AGO2 in ACC samples correlated with increased protein expression, further supporting its potential as a diagnostic biomarker for adrenocortical carcinoma.

### 3.2. Association between miRNA biogenesis gene expression and survival in ACC

To assess the prognostic value of miRNA biogenesis genes in Adrenocortical Carcinoma (ACC), we utilized RNA-seq data from The Cancer Genome Atlas (TCGA). For the survival analysis, cancer samples were divided into two groups based on the median expression of each gene, as per the guidelines provided by ENCORI. Among the genes involved in the miRNA biogenesis pathway, AGO2 emerged as the strongest prognostic indicator in ACC, exhibiting a hazard ratio (HR) of 7.07 and a Log-rank test p-value of 2.8e-06 (Fig 4). The Kaplan-Meier analysis further validated AGO2’s strong association with poor prognosis in ACC (Fig 5). Other genes such as DGCR8, XPO5, and RAN also demonstrated prognostic potential, but to a lesser extent, with HRs of 5.9 (p<0.0001), 4.25 (p=0.0004), and 5.06 (p=0.0001) respectively. TARBP2 showed a weaker prognostic association with a HR of 2.82 (p=0.014). On the other hand, DROSHA and DICER did not exhibit significant prognostic correlations, with HRs of 0.93 (p=0.85) and 1.24 (p=0.57) respectively.

**Figure 4:**
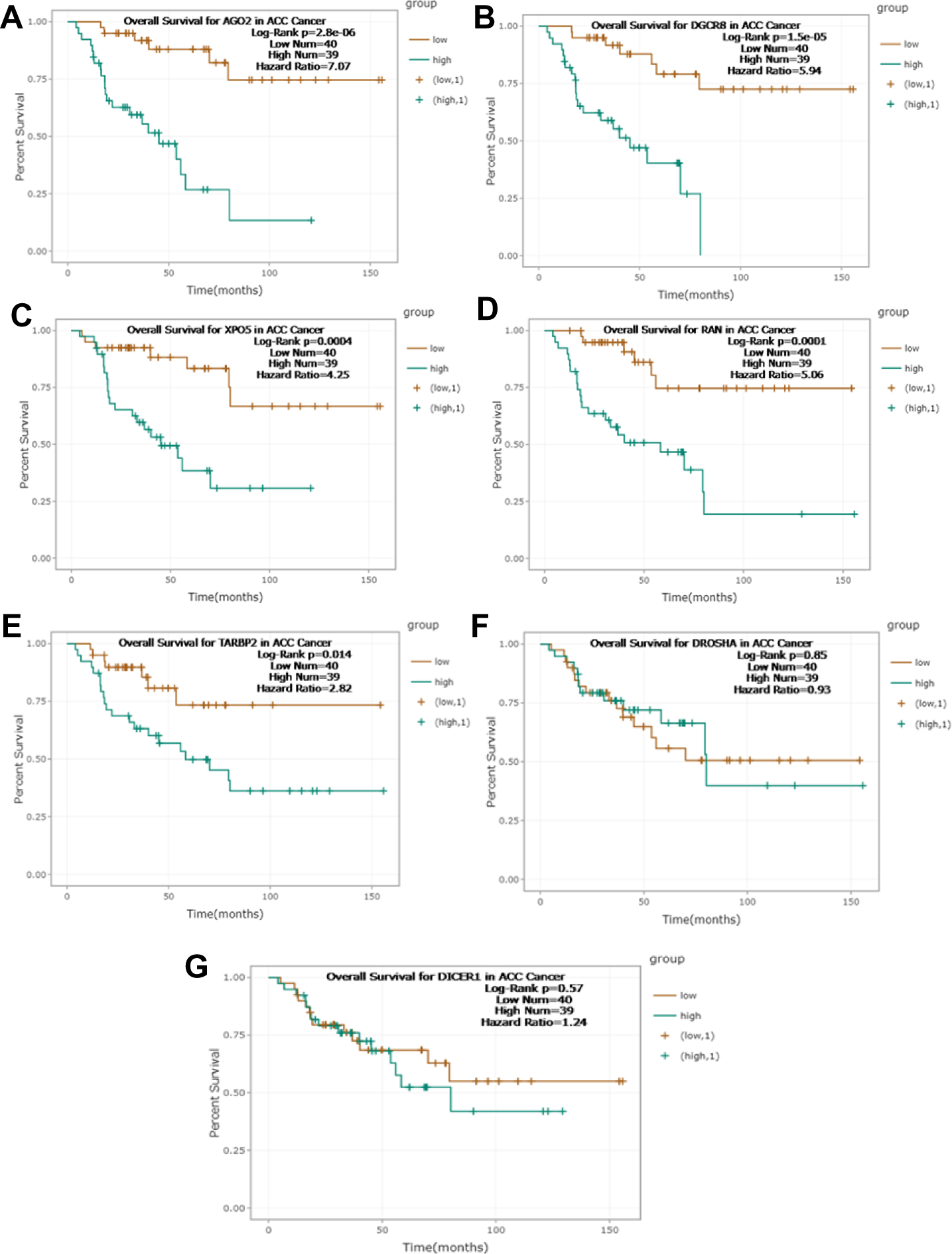
Association between miRNA biogenesis gene expression and survival rates in Adrenocortical Carcinoma (ACC). Gene Survival Analysis of TCGA RNA-seq data was performed to explore overall survival rates in 79 ACC patients with Adrenocortical Carcinoma according to high (green) or low (brown) gene expression levels. The analysis revealed a poor prognosis associated with high expression levels of AGO2, DGCR8, XPO5 and RAN with Log-Rank p <0.001. TARBP2 showed a weaker prognostic association with Log-Rank p=0.014. DROSHA and DICER did not exhibit significant prognostic correlations, with Log-Rank p=0.85 and p=0.57 respectively. Among the genes involved in the miRNA biogenesis pathway, AGO2 emerged as the strongest prognostic indicator in ACC, exhibiting a hazard ratio (HR) of 7.07 and a Log-rank test p-value of 2.8e-06 (35).

**Figure 5:**
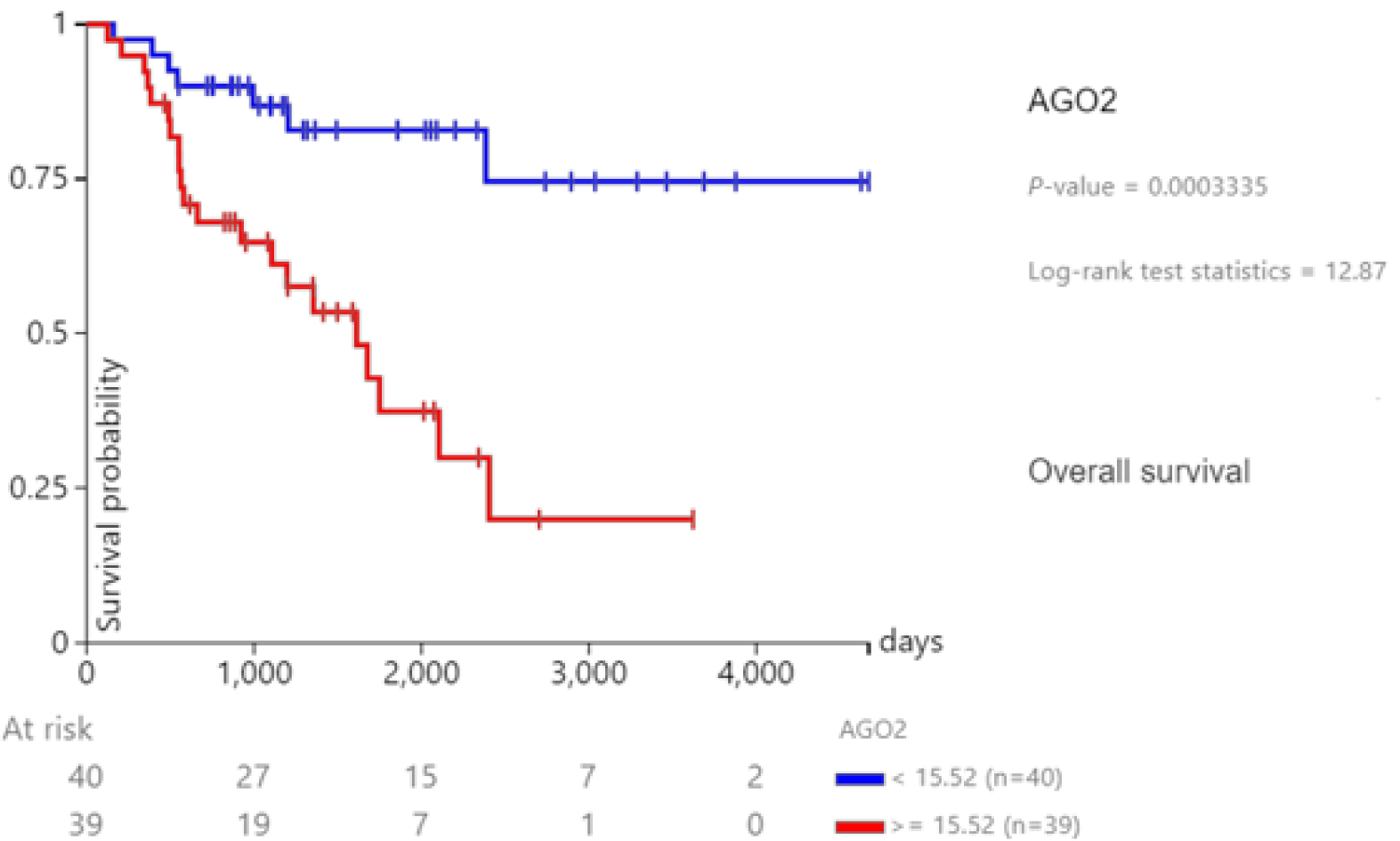
Kaplan Meier gene expression AGO2- ACC-TCGA. Kaplan-Meier curves compare survival between ACC patients with low (< 15.52, blue) and high (≥ 15.52, red) AGO2 expression in the TCGA cohort. The difference in survival is statistically significant (p = 0.0003335, log-rank test statistic = 12.87), indicating a prognostic impact of AGO2 expression on patient outcome (36).

### 3.3. Prognostic significance of AGO2 Gene Expression in ACC compared to other cancers

The prognostic correlation of AGO2 gene expression was strongest in ACC (HR 7.07, p=2.8e-06) compared to the 32 other TCGA cancer types studied (Figure 3). Although AGO2 gene expression held prognostic relevance in cholangiocarcinoma (HR 0.38, p=0.044), renal cell carcinoma (HR 2.15, p=0.016), mesothelioma (HR 2.36, p=0.00053), sarcoma (HR 1.71, p=0.0092) and endometrial carcinoma (HR 1.83, p=0.0052), in none of these other cancer did AGO2 demonstrate such a significant prognostic impact as in ACC (Fig 6, Table 1).

**Figure 6:**
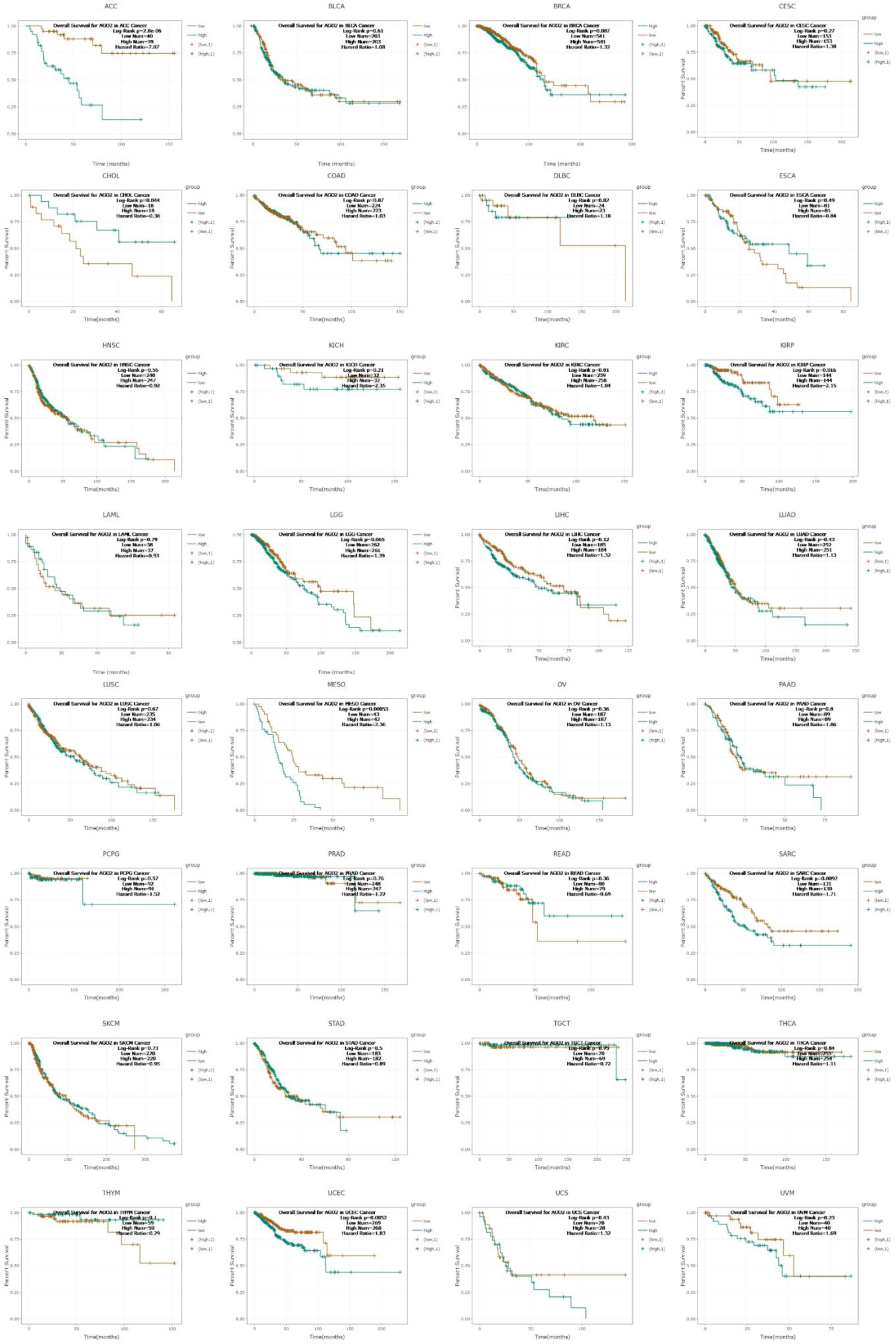
Association between AGO2 Expression and Patient Survival in Adrenocortical Carcinoma (ACC) and Other Cancer Types. To assess the prognostic value of AGO2 expression, gene survival analysis was performed using TCGA dataset. The analysis included 32 different cancer types, including ACC. The results revealed a significant association between differential expression of AGO2 and poor patient survival, specifically in ACC (p < 0.001). These findings underscore the significance of AGO2 expression as a prognostic marker for ACC, demonstrating its ability to predict patient survival. Furthermore, the analysis revealed the high significance of AGO2 as a prognostic marker in ACC compared with other cancer types in the TCGA dataset (35). See supplementary figure 6 for individual cancer type. The TCGA codes and their corresponding cancer types included ACC (Adrenocortical carcinoma), BLCA (Bladder urothelial carcinoma), BRCA (Breast invasive carcinoma), CESC (Cervical squamous cell carcinoma and endocervical adenocarcinoma), CHOL (Cholangiocarcinoma), COAD (Colon adenocarcinoma), DLBC (Lymphoid neoplasm diffuse large B-cell lymphoma), ESCA (Esophageal carcinoma), GBM (Glioblastoma multiforme), HNSC (Head and neck squamous cell carcinoma), KICH (Kidney chromophobe), KIRC (Kidney renal clear cell carcinoma), KIRP (Kidney renal papillary cell carcinoma), LAML (Acute myeloid leukemia), LGG (Brain lower grade glioma), LIHC (Liver hepatocellular carcinoma), LUAD (Lung adenocarcinoma), LUSC (Lung squamous cell carcinoma), OV (Ovarian serous cystadenocarcinoma), PAAD (Pancreatic adenocarcinoma), PCPG (Pheochromocytoma and paraganglioma), PRAD (Prostate adenocarcinoma), READ (Rectum adenocarcinoma), SARC (Sarcoma), SKCM (Skin cutaneous melanoma), STAD (Stomach adenocarcinoma), TGCT (Testicular germ cell tumours), THCA (Thyroid carcinoma), THYM (Thymoma), UCEC (Uterine corpus endometrial carcinoma), UCS (Uterine carcinosarcoma), and UVM (Uveal melanoma).

**Table 1.** Pan-Cancer AGO2 expression and survival analysis in TCGA cohorts. This table summarizes the hazard ratios (HR) for AGO2 expression across 32 TCGA cancer types, highlighting its prognostic significance, particularly in ACC with a HR of 7.07 (p value 2.80E-06) (35).

| Cancer | Cancer Number | p-value (significant threshold <0.05) | HR |
| --- | --- | --- | --- |
| ACC | 79 | 2.80E-06 | 7.07 |
| MESO | 85 | 0.00053 | 2.36 |
| UCEC | 537 | 0.0052 | 1.83 |
| SARC | 261 | 0.0092 | 1.71 |
| KIRP | 288 | 0.016 | 2.15 |
| CHOL | 36 | 0.044 | 0.38 |
| LGG | 523 | 0.065 | 1.39 |
| BRCA | 1082 | 0.087 | 1.32 |
| THYM | 118 | 0.1 | 0.29 |
| LIHC | 369 | 0.12 | 1.32 |
| KICH | 64 | 0.21 | 2.35 |
| UVM | 80 | 0.23 | 1.69 |
| CESC | 306 | 0.27 | 1.3 |
| OV | 374 | 0.36 | 1.13 |
| READ | 159 | 0.36 | 0.69 |
| LUAD | 503 | 0.43 | 1.13 |
| UCS | 56 | 0.43 | 1.32 |
| ESCA | 162 | 0.49 | 0.84 |
| STAD | 365 | 0.5 | 0.89 |
| HNSC | 495 | 0.56 | 0.92 |
| PCPG | 183 | 0.57 | 1.52 |
| BLCA | 406 | 0.61 | 1.08 |
| LUSC | 469 | 0.67 | 1.06 |
| SKCM | 440 | 0.73 | 0.95 |
| TGCT | 139 | 0.75 | 0.72 |
| PRAD | 495 | 0.76 | 1.22 |
| LAML | 75 | 0.79 | 0.93 |
| PAAD | 178 | 0.8 | 1.06 |
| KIRC | 517 | 0.81 | 1.04 |
| DLBC | 47 | 0.82 | 1.18 |
| THCA | 509 | 0.84 | 1.11 |
| COAD | 447 | 0.87 | 1.03 |

### 3.4. Prognostic implications of AGO2 gene expression in cancer staging

Using TCGA ACC data in the Xena browser, a one-way ANOVA revealed a significant correlation between tumour stage and the gene expression of AGO2 (p = 0.038) and RAN (p = 0.013) in adrenocortical carcinoma. Other genes, including DGCR8, DROSHA, DICER1, TARBP2, and XPO5, did not show significant associations (Fig 7).

**Figure 7:**
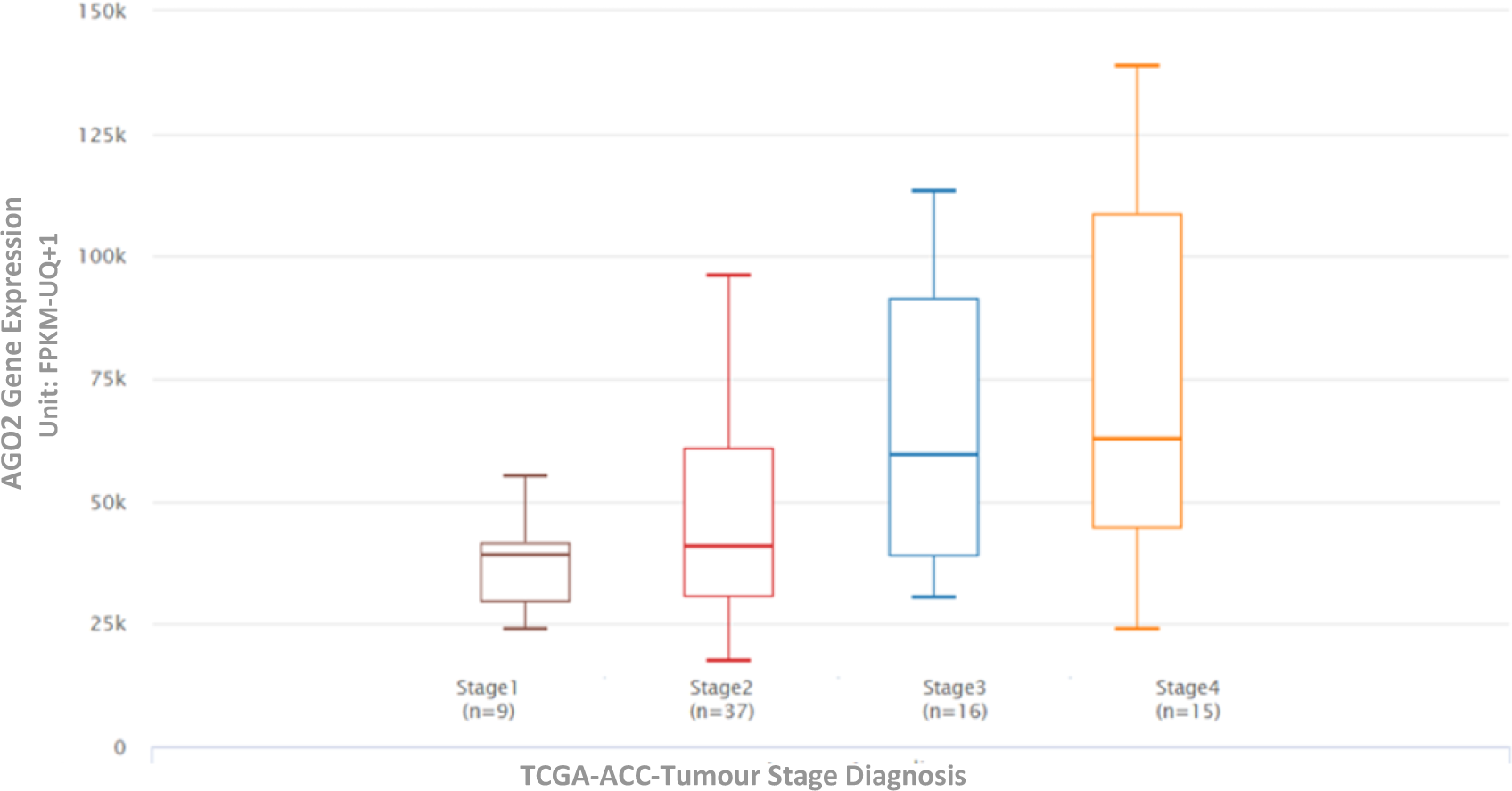
Prognostic significance of AGO2 expression in Adrenocortical Carcinoma (ACC) patients based on cancer staging: The expression levels of AGO2 were compared among 79 TCGA ACC patients across different cancer stages. The figure shows that the gene expression of AGO2 was highest in Stage IV patients compared to that in other stages. This suggests that AGO2 expression may play a role in disease progression and could be a potential prognostic marker for ACC. Overall, the results shown in Figure 6 suggest that AGO2 expression can be used in conjunction with cancer staging to predict the prognostic outcomes of ACC. Specifically, high AGO2 expression in stage IV patients is associated with poor patient survival. (36)

### 3.5. Differential protein expression patterns of miRNA biogenesis genes in ACC, adrenal adenoma and normal adrenal cortex

AGO2 protein concentration was significantly higher in ACC than in adrenal adenoma or normal adrenal cortex (p<0.0001). Furthermore, there was no significant difference in AGO2 protein expression between normal and benign tumour (Figure 8). In contrast, XPO5, RAN, and DICER1 protein expression levels were significantly lower in ACC tissue samples compared to the non-malignant groups (p < 0.001). No statistically significant differences were observed in the protein expression levels of DROSHA, DGCR8, or TARBP2 between the malignant and non-malignant groups.

**Figure 8:**
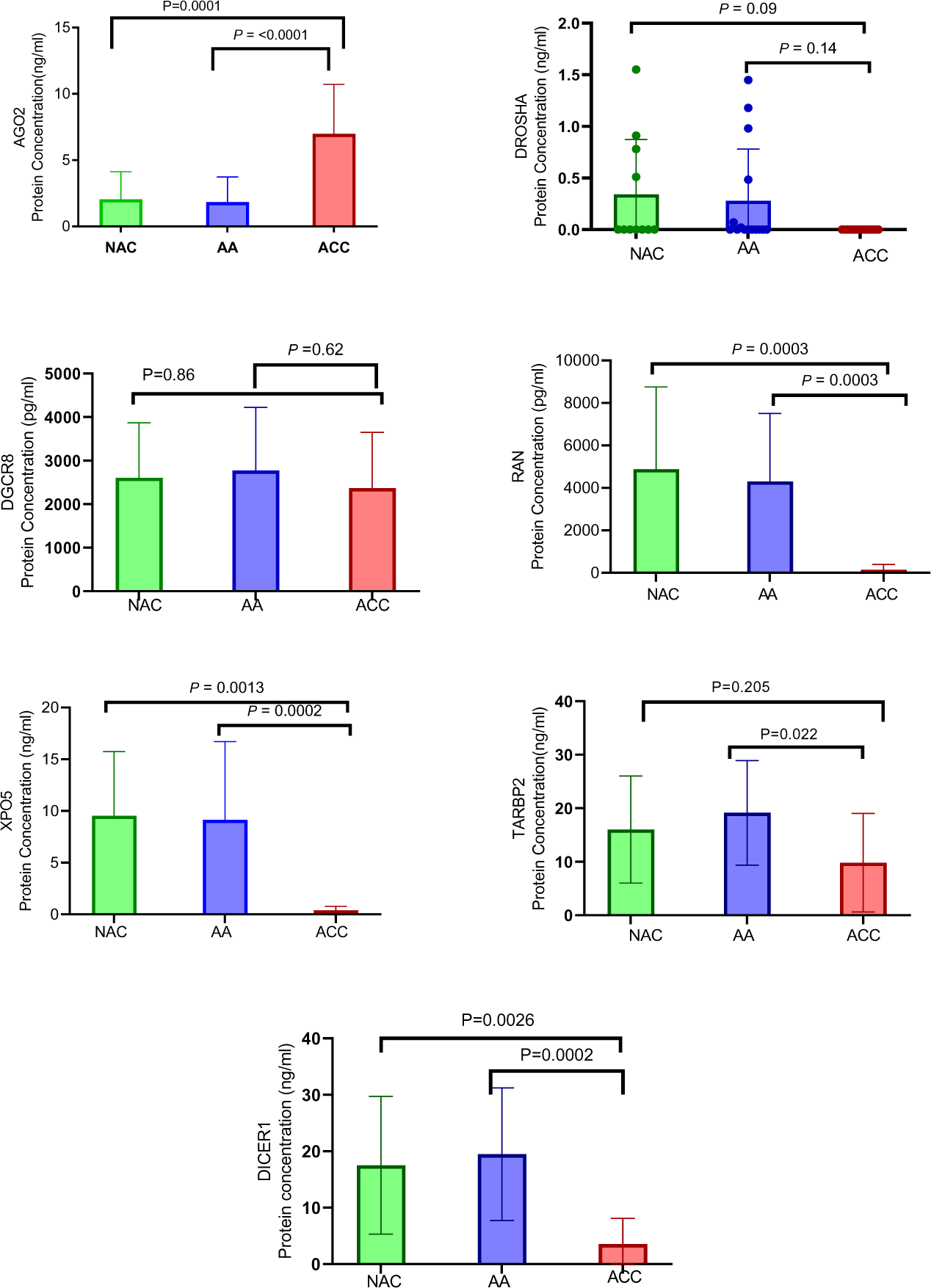
Protein Expression Analysis of miRNA Biogenesis Genes in Adrenocortical Carcinoma (ACC), Normal, and Benign Tissue Samples. Protein expression levels of DROSHA, DGCR8, XPO5, RAN, TARBP2, DICER1, and AGO2 were measured using ELISA in normal, benign, and adrenocortical cancer tissue samples. The results revealed that XPO5, RAN, TARBP2, and DICER1 protein expression was downregulated in cancer samples compared to that in both normal and benign samples, suggesting a potential role of these proteins in cancer development through post-translational modification. In contrast, AGO2 showed significantly higher protein expression in cancer samples than in normal samples (p<0.001), and its protein expression was also higher in cancer samples than in both normal and benign samples. These findings highlight AGO2 as a strong candidate for a potential diagnostic biomarker for adrenocortical carcinoma among all the miRNA biogenesis factors analyzed.

#### 3.5.1. ROC analysis, specific cut-point determination

To explore the appropriate diagnostic threshold of AGO2 protein expression levels in ACC compared to non-malignant tissue, we performed a Receiver Operating Characteristic (ROC) curve analysis. The area under the curve (AUC) was 0.95 (95% CI: 0.86 to 1.00), indicating a high diagnostic accuracy. Using a cut-point of >3.9ng/ml for AGO2 protein expression, a sensitivity of 89% (95% CI: 57% to 99%) and a specificity of 80% (95% CI: 55% to 93%) was achieved (Fig 9).

**Figure 9:**
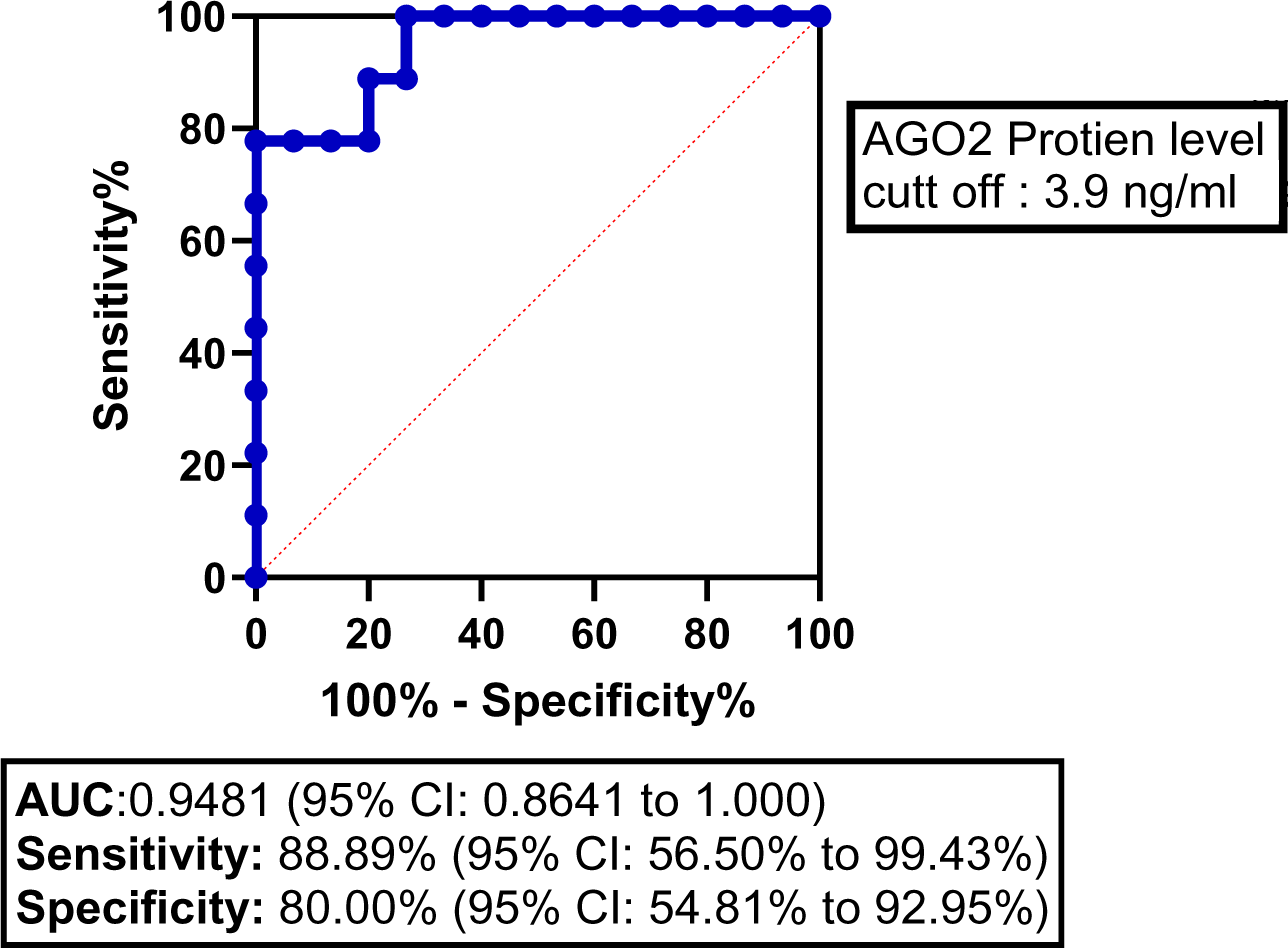
Receiver Operating Characteristic (ROC) Curve for AGO2 Protein Expression in Adrenocortical Carcinoma (ACC). The ROC curve illustrates the diagnostic ability of AGO2 protein expression to differentiate between ACC and non-malignant samples. The area under the curve (AUC) is 0.9481 (95% CI: 0.8641 to 1.000), indicating a high diagnostic accuracy. A cut-off value of >3.9 for AGO2 protein expression yielded a sensitivity of 88.89% (95% CI: 56.50% to 99.43%) and a specificity of 80.00% (95% CI: 54.81% to 92.95%). The diagonal dashed line represents the line of no discrimination (AUC = 0.5).

## 4. Prognostic significance of AGO2 protein expression in relation to clinicopathological characteristics in TCGA-ACC patients

The prognostic potential of AGO2 in ACC was further explored by examining its association with various clinicopathological characteristics. Table 2 presents a detailed analysis of five ACC patients from the TCGA cohort, for whom a protein expression analysis was conducted using samples retrieved from the Kolling Tumour Bank. Higher AGO2 protein expression correlated with advanced tumour stage, higher Weiss histopathological scoring, Ki67 proliferative index and poorer survival outcomes.

**Table 2:** Prognostic significance of AGO2 protein expression in relation to clinicopathological characteristics in TCGA-ACC patients.

| Sample | AGO2 Protein (ng/ml) | Weiss Score | Tumour Stage | Ki67 Index% | Gender | Vital Status | CT-Scan findings |
| --- | --- | --- | --- | --- | --- | --- | --- |
| TCGA-8OR-A5JJ-01A | 14.31 | 9 | iv | 30 | M | Dead | Lung metatasis |
| TCGA-1OR-A5JM-01A | 10.18 | 9 | iv | unknown | F | Dead | Liver metatasis |
| TCGA-9OR-A5JG-01A | 9 | 9 | iv | 40 | M | Dead | Retroperitoneal Lymph Node |
| TCGA-3OR-A5JK-01A | 4.02 | 8 | iv | 5 | M | Alive | Lung metastasis |
| TCGA-7OR-A5JF-01A | 3.64 | 4 | ii | 10 | F | Alive | Unknown |

## 5. Discussion

In this study, we demonstrate the potential role of AGO2 as a diagnostic and prognostic marker in ACC, with protein expression levels that stratify ACC from NAC and AA, as well as correlating with disease stage and prognosis. Of all the potential miRNA biogenesis proteins we examined, AGO2 has the greatest potential to be feasibly translatable to a clinical setting. For example, while TARBP2 and RAN also exhibited gene overexpression in ACC compared to the normal adrenal cortex, they did not show a corresponding increase at the protein level. Similarly, although XPO5 protein expression was significantly reduced in ACC relative to adenoma and normal adrenal cortex, it was high rather than low levels of XPO5 protein that were associated with adverse prognostic outcomes on survival analysis. Conversely, AGO2 demonstrated concordance between gene expression, protein expression and prognostic impact on survival analysis results, as well as demonstrating the highest adversely prognostic hazard ratio.

Our analysis of mRNA sequencing data comparing ACC to the 32 cancer types in the TCGA revealed that AGO2 expression was by far the most prognostically important in ACC. This may indicate a significant role of AGO2 in ACC pathogenesis and is a finding that warrants further exploration. Irrespective of pathogenic mechanism, our results indicate that AGO2 may be a viable diagnostic and prognostic biomarker in clinical practice. Previous studies that have examined the prognostic impact of miRNA biogenesis proteins have reported conflicting results. For example, Carmuta (31) reported upregulation of TARBP2 mRNA levels in ACC patients, whereas de Sousa (32) found no difference in TARBP2 gene or protein (TRBP) expression between adrenocortical adenomas and ACC. In our study, although TARBP2 and RAN gene expression was significantly increased in ACC, a corresponding increase in protein expression was not seen. Conversely, the gene expression of DROSHA, XPO5, and DICER did not differ between ACC and normal adrenal cortex, however the levels of protein expression were significantly lower in ACC. These discrepancies not only highlight the complexity of post-transcriptional and post-translational regulatory mechanisms on protein expression levels in ACC, as well as the inherent challenges in comparing different methodological quantitative approaches.

It is well established that miRNAs play a critical role in tumorigenesis (38,39)and AGO2 plays as a key role in regulating miRNAs function and maturation.(40) Although AGO2 overexpression has been documented in several carcinomas, including colon cancer, head and neck squamous cell cancer, urothelial carcinoma of the bladder, ovarian carcinoma, gastric carcinoma, and colorectal carcinoma (41), the role of AGO2 is not uniform across all cancer types. For instance, in melanoma, AGO2 expression is notably reduced at the protein level, despite stable mRNA levels. Intriguingly, over-expression of AGO2 in this context actually inhibits cell and tumour growth(42). This contradiction suggests that AGO2’s expression and its downstream effects may differ between cancer types, potentially due to distinct miRNAs expression patterns. Further research is required to understand the subtleties underlying the variance in AGO2 expression and its oncogenic effect.

In progressing toward clinical translation, several considerations must be addressed. Establishing the cut-point for AGO2 protein expression is important. Furthermore, comparison of AGO2 protein levels in tissue samples and blood samples may facilitate further investigation into its potential application as a liquid biopsy. Similarly further investigation into the quantitative significance of AGO2 protein levels in early-stage tumours may be useful in guiding adjuvant treatment and follow-up protocols.

## 6. Limitations

This study is limited by small sample size, and further validation in a larger cohort is required. Additionally, the establishment of clinically relevant cut-off values for AGO2 protein expression in tissue samples requires additional clinical trials and validation in larger cohorts.

## 7. Conclusion

AGO2 is upregulated in ACC in comparison to adrenal adenoma and normal adrenal cortex. This upregulation was evident at both the gene and protein levels. In comparison to the 32 other cancer in the TCGA dataset, the degree and significance of prognostic impact of AGO2 expression was unique to ACC. The strong association between AGO2 expression and clinicopathological outcomes highlights its potential role as a diagnostic and prognostic biomarker in ACC.

This study lay the groundwork for future research, particularly in exploring the feasibility of AGO2 as a liquid biopsy biomarker, a promising avenue that could revolutionize non-invasive cancer diagnostics and prognostication in ACC.

## Data Availability

All data produced in the present study are available upon reasonable request to the authors

